# Peri-tumoural lymphocyte neighbourhoods predict longer survival in pancreatic ductal adenocarcinoma

**DOI:** 10.1101/2025.10.14.25338000

**Authors:** Riley J. Arseneau, Jorge P. Mejia, Sarah Nersesian, Stacey N. Lee, Carley Bekkers, Thomas Samson, Daniel Gaston, Boris L. Gala-Lopez, Ravi Ramjeesingh, Thomas Arnason, Jeanette E. Boudreau

## Abstract

**Background:** Pancreatic ductal adenocarcinoma (PDAC) remains one of the deadliest cancers, and has limited options for treatment. Low immune infiltration, desmoplastic stroma, and poor tumour immunogenicity are all expected to contribute to PDAC’s rapid progression and limited response to existing immunotherapies. PDAC tumours are mosaics of different sub-tumour microenvironments, including some that do contain immune cells. We hypothesized that increasing frequency of lymphocyte:tumour interactions would correlate with lengthened survival for patients with PDAC.

**Methods:** Using multiplex immunofluorescence, digital pathology, and computational analyses, we profiled the spatial distribution, co-localization, and neighbourhood architecture of immune cells in tumours from 73 patients with PDAC.

**Results:** Higher densities of CD3+CD8– (CD4+) T cells were associated with improved five-year overall survival, particularly when enriched at the tumour-stroma boundary (i.e. peritumoural). CD3+CD8-T cells, CD8+ T cells (CD3+CD8+), and B cells (CD20+) frequently co-infiltrated and co-localized, forming distinct immune neighbourhoods indicative of organized adaptive immunity. We defined three common immune neighbourhoods within PDAC (1) Macrophage-dominant; (2) T cell dominant; and (3) Disorganized. With greater tumour and peri-tumoural area represented by the T cell dominant neighbourhood, overall survival was increased. The other neighbourhoods were not significantly associated with outcomes.

**Conclusion:** Immune cells self-assemble in predictable patterns in PDAC. The presence of T cell dominant neighbourhoods, which we interpret as supporting ongoing immune activation, best predict lengthened survival. Spatially organized immune interactions may serve as prognostic indicators and inform future studies for immunotherapies in PDAC.

## Introduction

Pancreatic ductal adenocarcinoma (PDAC) remains fatal in greater than 80% of patients, with few options for effective treatment.^1^ immunotherapies like immune checkpoint blockade or chimeric antigen receptor (CAR)-T cells have transformed treatment of other cancers, such as melanoma and leukemia,^2,3^ but PDAC remains refractory to existing immunotherapeutic interventions.^4^ PDAC is often classified as an immunologically “cold” tumour, with a tumour microenvironment (TME) marked by dense desmoplasia, poor immune cell infiltration, a low mutational burden/neoantigen availability, and a high density of immunosuppressive cells such as M2 macrophages and regulatory T cells (Tregs).^4,5^ However, recent immunohistochemical assessments have revealed that PDAC is heterogenous both within and between patients, a product of its “sub-TMEs”.^6^ Among these are immune-enriched regions; this implies that immunity (and immunotherapy) remain possible in PDAC.^6–10^ In agreement, recent clinical investigations have demonstrated that neoantigen-targeting personalized vaccines can drive effective cytotoxic T cell anti-tumour immunity in patients with PDAC^11,12^. There is thus a need to better understand the immune microenvironment in PDAC: the composition of tumour and immune cells and their spatial interactions may provide prognostic insights that ultimately inform more precise care and future immunotherapies.

Immune-mediated tumour killing requires contact between tumour cells and cytotoxic immune cells or their products (i.e. antibodies). Activation for anti-tumour immunity requires a co-ordinated effort between immune cells that requires contact between antigen presenting cells, helper T cells and effector lymphocytes. Increasingly, the spatial context of immune-tumour interactions is being interrogated to better understand putative cell-cell communication, activation, and priming—and predicting patient outcomes.^13–17^ For example, tertiary lymphoid structures that localize T cells, B cells, and antigen presenting cells enable anticancer immunity.^18^ We recently demonstrated that clusters of macrophages, T cells, and natural killer (NK) cells infiltrating the tumour epithelial regions predicted the longest progression free survival in patients with high grade serous carcinoma of the ovary.^16^ The co-localization of immune cells with complementary functions to collaboratively drive anticancer immunity best associated with patient survival, perhaps underscoring a requirement for immune cell co-operation in tumour microenvironments.^16^ Whether a similar co-operation of immunity occurs in PDAC has not been investigated.

In this study, we used multiplex immunofluorescence (mIF) to assess the spatial distribution, co-localization, and neighbourhood formation of macrophages, T cells, NK cells, and B cells in 73 surgically-resected PDAC tumours. We designed a novel approach to segregate tumours into epithelial, peri-tumoural and stromal regions, because unlike most solid tumours where there is a clear delineation of tumour epithelium from its surrounding stroma, PDAC initiates in ducts and metastasizes via small satellite tumour clusters.^19,20^ We observed that enrichment of CD3+CD8– T cells (likely CD4+ helper T cells) was associated with longer five-year overall survival (OS), particularly when they are at the tumour-stroma (peritumoural) interface. The survival benefit was further enhanced in patients with CD3+CD8-T cells in the stroma co-occurring with CD3+CD8+ (cytotoxic) T cells in the tumour epithelium, consistent with CD8+ T cell activation supported by regional support from helper T cells. Unsupervised clustering revealed three patterns of immune cell co-localization (“neighbourhoods”); one among them, containing CD3+CD8– T cells, CD3+CD8+ T cells, and B cells, was associated with the best outcomes for patients. This neighbourhood may represent regions of organized adaptive immunity and their presence in PDAC could therefore represent a productive anti-cancer response. Hence, these T cell-enriched areas may inform patient prognosis or precision medicine. While functional assessments will be required to determine the specific anti-cancer activities and driving features of these neighbourhoods, their presence underscores optimism for future, novel immunotherapies to support cytotoxic immunity in PDAC.

## Methods

### Patient samples

Samples were obtained from 76 patients with PDAC who underwent surgical resection from 2013 to 2024 at the Queen Elizabeth II Health Sciences Centre (Nova Scotia, CAN). This study was approved by the Nova Scotia Health institutional research ethics board (#1023467 & #1026628). Patients who received neoadjuvant therapy were excluded. Patient information is available in **Supplementary Table 1.**

Patient samples were formalin-fixed, paraffin-embedded (FFPE) and assembled into tissue microarrays (TMA). Each patient was represented by duplicate 3 mm cores selected from regions containing >30% tumour cells, as identified by board-certified pathologist (TA).

### Multiplex immunofluorescence

Opal mIF (Akoya Biotechnology, Marlborough, USA) was performed as previously described.^16^ Briefly, 4 µm sections were prepared from the TMAs, deparaffinized in Xylene (Sigma-Aldrich) (3 × 10 min) and rehydrated by incubating with decreasing concentrations of ethanol for 10 min each (100%, 100%, 95%, 70%). The slides were then fixed in 10% neutral buffered formalin (VWR) for 20 min.

The slides underwent serial cycles of antigen retrieval, primary staining, and tyramide signal amplification to add each antibody and fluorophore one by one. For antigen retrieval, slides were microwaved in Tris-EDTA buffer (pH9) at 1000W for 2 min, then 200W for 15 min. The slides were cooled on ice for 10 min and rinsed in deionized water and Tris-buffered saline with Tween-20 (TBS-T). Slides were thereafter blocked in 1x antibody diluent/block (Akoya Biosciences) for 10 min, the excess block was removed, and the slides were stained with diluted primary antibody for 45 min. After TBS-T rinsing, slides were incubated in secondary anti-mouse plus rabbit antibodies conjugated to horseradish peroxidase (HRP) for 10 min. Signal amplification was completed via HRP-driven tyramide activation and fluorophore deposition and tissue binding (Opal 6-Plex Manual Detection Kit, and Opal 480 Reagent Pack (Akoya Biosciences)). Opal 780 staining (Opal 780 Reagent Pack, Akoya Biosciences) required use of digoxygenin labeling prior to tyramide signal amplification (10 min, TSA Plus DIG, Akoya Biosciences). The slides were rinsed in TBS-T and subsequently incubated in the corresponding Opal fluorophore diluted in Opal amplification diluent for 10 min (Opal 480, 520, 540, 570, 620, 650, and 690 fluorophores) or 1h (Opal 780). After the final staining step, slides were counterstained with Spectral DAPI (ThermoFisher) for 10 min, rinsed, and mounted in Mowiol-based mounting medium (Mowiol, glycerol, deionized H2O, TrisHCl (pH 8.5); Sigma-Aldrich)). Antibodies, Opal fluorophores with which they were used, and staining order are shown in **Table 1**.

**Table 1.** Staining panel and order for mIF.

| Table 1. Staining panel and order for mIF |  |  |  |  |
| --- | --- | --- | --- | --- |
| Phenotype | Marker | Clone | Dilution | Fluorophore |
| NK cells | CD57 | NK1 | 1/50 | 650 |
| M2 Macrophage | CD163 | ERP19518 | 1/200 | 540 |
| Macrophage/monocyte | CD68 | SP251 | 1/100 | 690 |
| Epithelial Cell | PanCk | AE1/AE3 | 1/100 | 520 |
| Pan T cell | CD3 | LN10 | 1/50 | 480 |
| NK cells/monocyte | CD16a | SP175 | 1/50 | 620 |
| Cytotoxic T cell | CD8 | C8/144 | 1x | 480 |
| B cell | CD20 | L26 | 1/25 | 780 |
| DAPI (nuclear stain) | DAPI | N/A | 15 μM | N/A |

Antibody staining was validated using FFPE tonsil tissue and cell pellets of purified T and NK cells. T and NK cells were isolated from primary peripheral blood mononuclear cells (Canadian Blood Services, Dalhousie REB 2016-3842), centrifuged at 300 x *g* for 5 min, fixed in 10% neutral buffered formalin, and processed for paraffin embedding. All staining patterns were confirmed by a board-certified pathologist (TA). Representative images from one case and the segmentation pipeline are shown in **Figure 1A**.

**Figure 1.**
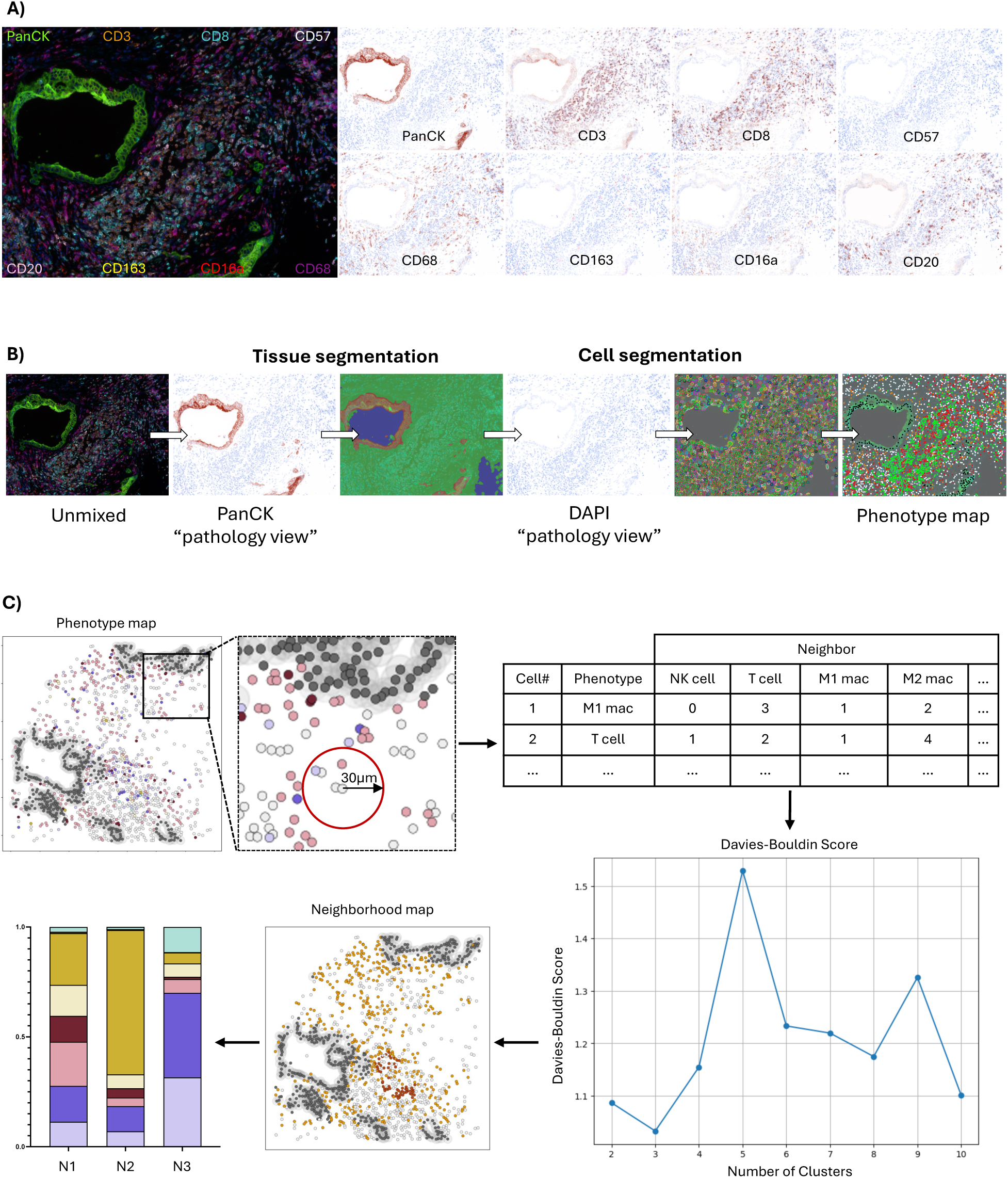
Quantitative image and neighbourhood analysis workflows. **A)** Fluorescent microscopy image using 9-colour mIF. Markers used include DAPI (blue), CD3 (orange), CD8 (cyan), CD57 (white), CD16a (red), CD68 (magenta), CD163 (yellow), CD20 (pink), and panCK (green). Representative pathology views for each marker are shown. **B)** Quantitative pathology workflow with tissue and cell segmentation based on PanCK and DAPI staining, respectively. **C)** x,y coordinates were used to reconstruct maps of tumours *in silico*. Cell groups were defined as at least five cells within a 30 µm radius and k-means clustering identified cell neighbourhoods. Davies-Bouldin indexing was used to define the ideal number of neighbourhoods, whose cellular compositions are reported as bar graphs.

Imaging was completed using a Mantra Quantitative Pathology workstation equipped with after-market Opal 480 and Opal 780 filter cubes. Images were acquired at 20x magnification, with wavelength intervals spanning emission peaks between 420 nm and 780 nm to capture all Opal fluorophores. Up to 15 images per core were captured, with multispectral analysis performed in InForm Tissue Finder Software (Version 2.4.8). Spectral reference libraries, generated from monoplex stains for each fluorophore, were used for spectral unmixing to distinguish fluorophore signals and control for autofluorescence. Regions of benign panCK+ staining were omitted and regions of tumour epithelium were confirmed by a board-certified pathologist (TA).

Using machine learning algorithms, tissue segmentation was performed on spectrally unmixed images to distinguish epithelial and stromal regions (off-core regions were omitted and could be distinguished from stroma by lack of DAPI staining and tissue autofluorescence). Nuclear segmentation (DAPI) followed, and cell phenotypes were assigned by antibody staining. Phenotype maps with x,y cell coordinates classified each cell with a unique cell ID **(Figure 1B)**. Cell types were classified based on antibody expression profiles: panCK+ as epithelial cells (regardless of other antibody binding), PanCK-CD68+ as macrophages (CD163+/–, CD16a+/–), PanCK–CD68–CD3+ as T cells (CD8+/–, CD16a+/–, CD57+/–), PanCK–CD3–CD68–CD57+ as NK cells (CD16a+/–), PanCK– CD3–CD68–CD57–CD16a+ as CD16a+ cells, and panCK–CD3–CD68–CD20+ as B cells. Cells staining with DAPI only were categorized as “Other”. Antibody binding patterns, cell surface staining, cell segmentation, and phenotyping were validated by a board-certified pathologist (TA).

Quality control involved visual assessment of every image and excluding images with poor tissue quality or non-specific staining. All quality control was performed blinded to clinical outcome data. Of the 76 patients intended for study, images from 73 patients passed staining and quality control and were included in the final analysis.

### Cell densities and spatial analyses

#### Cell densities

Cell identities and x,y coordinates were exported from InForm. Cell densities were calculated in RStudio (Boston, MA USA, version 2022.12.0) using the dplyr package.^21^ For each patient, total counts of each cell type between duplicate cores within the epithelial and stromal compartments were summed and divided by the corresponding tissue area, resulting in region-normalized average densities per patient (cells per mm^2^). For cell-, neighbourhood, and region-level analyses, we calculated the density (cells/mm^2^) of each immune cell subset (or neighbourhood) per patient. Density distribution curves were generated using the density () function in base R, and the division into “high” and “low” densities was defined by the maxima, minima, or inflection point closest to the median of each distribution.

#### T cell infiltration score

We generated a T cell infiltration score, following an approach like that used in Immunocore algorithm used in colorectal cancer.^22,23^ The T cell infiltration score ranged from 0 to 6 and was calculated by summing binary high/low infiltration designations of CD3+CD8+ and CD3+CD8– T cells across the epithelial and stroma compartments. Informed by compartment specific role of each subset,^24–27^ we assigned a greater weight to the subset most biologically relevant in each region. Patients received two points for high CD3+CD8– T cell infiltration in the stroma and one point for high CD3+CD8+ infiltration in the stroma. Conversely, high CD3+CD8+ infiltration in the epithelium contributed two points, while CD3+CD8– in the epithelium contributed one point. Patients were stratified into three groups based on total score: low (0 points), intermediate (1-3 points), and high (4-6 points).

#### Neighbourhood analysis

Our spatial biology workflow was adapted from previously published methods^15,16^ and conducted in Python (Version 3.9.18). X,y coordinates of each cell were used to reconstruct phenotype maps of tumours, and each cell was defined as an index. Cell IDs with their x,y coordinates and assigned phenotypes were grouped by image and patient using Pandas^28^ (for data organization and manipulation) and NumPy^29^ (for numerical operations). Using the BallTree module from scikit-learn^30^, for each index cell, cells within a 30 µm radius were identified based on their x,y, coordinates (nearest neighbour search), defining local “groups” **(Figure 1C)**. To identify patterns of immune cell co-localization, which we termed “neighbourhoods”, three images from each patient were randomly selected and combined to create a training set. Using this set, K-means clustering through MiniBatchKMeans algorithm (scikit-learn) was applied to the immune cell populations only, testing between 2 and 10 potential neighbourhoods. The optimal number of neighbourhoods to represent intra-tumour diversity was defined using Davies-Bouldin indexing (scikit-learn).^31^

All remaining images were classified using the trained model to assign each cell to a neighbourhood. To ensure neighborhood definitions reflected immune cell organization (not tumour or other cells), epithelial (PanCK+) and “Other” cells were excluded from the clustering and instead assigned to distinct “PanCK” and “Other” neighborhoods. This approach preserved their spatial position in the radius-based analysis while preventing them from influencing immune neighbourhood structure. Consistency of neighbourhood classification was validated by calculating cosine similarity (scikit-learn), which scored >0.99 between the initial training set and the subsequent classified images. Figures were generated with matplotlib^32^ and seaborn^33^.

#### Tumour neighbourhood analysis

To define tumour regions as intratumoural, peritumoural, and stromal, cell clustering was performed in Python using the DBSCAN^34^ and SciPy^35^ packages. Epithelial (panCK+) cells were first identified in each sample, and DBSCAN clustering was performed on these cells using a 30 μm radius and a minimum cluster size of five cells. Cells that did not meet this clustering criteria were excluded from defining intratumoural regions. SciPy’s ConvexHull class was used to define the smallest bounding shape (convex hull) for each tumour cluster, with visual confirmation by a pathologist (TA). Based on these boundaries, three regions were defined: (1) Intratumoural: the area formed by each tumour site; (2) Peritumoural: a 30 μm perimeter surrounding each tumour site; or (3) Stromal: the area outside the intratumoural and peritumoural regions. Cell counts for each region were aggregated by radius for each patient, and patients were subsequently classified into high and low groups based on the proportion of each immune phenotype within the defined regions, as defined above.

### Statistical analysis

To define which clinical variables were associated with survival and therefore should be used as covariates for downstream multivariate analysis, we performed univariate Cox proportional hazard regressions using five-year OS as the dependent variable in RStudio. Independent variables were tumour stage, tumour grade, age at diagnosis, sex, and lymph node invasion **(Supplemental Table S1)**. Among them, only increased regional lymph node metastasis was significantly associated with shorter five-year OS (p < 0.05).

Univariate Kaplan-Meier and log-rank tests were conducted to assess differences in OS between high and low groups of cell-, neighborhood-, and regional-analyses, and multivariate Cox proportional hazards models were used to identify independent prognostic factors. Co-infiltration between cellular phenotypes were assessed using Spearman’s rank correlation in GraphPad Prism (GraphPad Software, Boston, MA USA, version 10.2.3). Comparisons of immune cell infiltration across clinically defined groups, including tumour stage (T1, T2, T3), tumour grade (1, 2, 3), age (<65, ≥65), sex (male vs female), and regional lymph node metastasis (N0, N1, N2), were conducted using two-tailed Mann-Whiteney U tests in GraphPad Prism, with corrections for false discovery to account for multiple comparisons. Statistical significance was defined as an alpha of 0.05 for all analyses.

## Results

### Patient characteristics

Using Opal mIF, we evaluated tumours from a total of 73 patients with PDAC who underwent surgical resection between 2013 and 2024 at the QEII Health Sciences Centre. Forty-five percent of the cohort was female; 55% were male **(Supplemental Table 1)**. Most patients were >65 years old (59%, n=43), and categorized as stage II (33%) or stage III (62%). Moderately differentiated tumours were most common (63%, grade 2), followed by poorly differentiated (23%, grade 3), and well-differentiated tumours (14%, grade 1). Regional lymph node involvement was most commonly present: just 14% had no positive lymph nodes (N0), 60% of patients had 1-3 positive lymph nodes (N1), and 21% having ≥4 nodes (N2). Patients with N0 disease had a lower risk of death compared to N2 (HR: 0.16, 95% CI: 0.04-0.73, p<0.05). Since this was the only clinical variable associated with OS, it was the only one included in downstream multivariate analyses.

### Immune architecture of PDAC microenvironment

The immune microenvironment of PDAC is immunosuppressive and spatially heterogenous, yet remains poorly characterized, with limited knowledge of how it reflects patient prognosis.^4,5^ To characterize the immune landscape of PDAC, we quantified immune cell densities (cells/mm^2^) in both the epithelial and stromal compartments using multiplex IHC and quantitative pathology **(Figure 1)**. Tumours displayed marked immune infiltration with inter- and intra-patient heterogeneity **(Figure 2A)**. Regardless of phenotype, immune cell densities were higher in the stroma than in the epithelial compartment, likely reflecting limited accessibility to the tumour **(Figure 2A)**. Among immune populations, the most abundant were CD16a+ M2-like macrophages (epithelium median: 11.3, min-max: 0-280.1; stroma median: 569.1, min-max: 45.2-4602.0) and CD3+CD8+ T cells (epithelium median: 10.6, min-max: 0-216.1; stroma median: 234.4, min-max: 24.9-1446.0), reflecting substantial infiltration in both compartments **(Figure 2A, Supplemental Table 2).** To reduce complexity and improve interpretability, populations with a median cohort total density of <1 cell/mm^2^ were grouped into higher-level phenotypes based on shared marker expression **(Supplemental Table 2)**. Specifically, PanCK–CD68–CD3+CD8–CD57+ and PanCK–CD68– CD3+CD8–CD16a+ subsets were amalgamated into a CD3+CD8– T cell group; PanCK–CD68– CD3+CD8+CD57+ and PanCK–CD68–CD3+CD8+CD16a+ subsets were grouped as CD3+CD8+ T cells; and PanCK–CD68–CD3-CD8–CD57+CD16a– and PanCK–CD68–CD3-CD57+CD16a+ subsets were grouped as NK cells.

**Figure 2.**
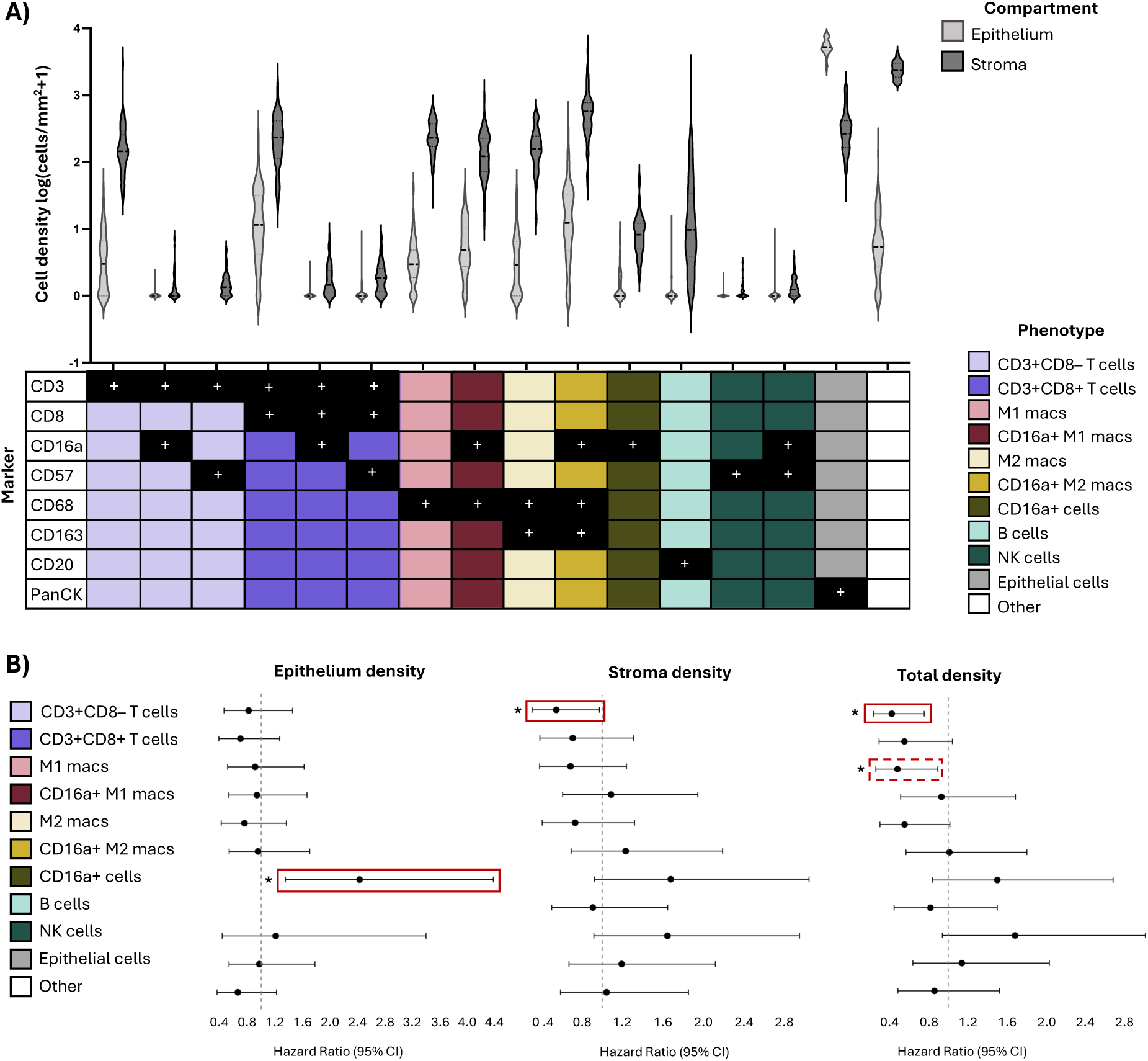
Immune cell frequencies and associations with survival. **(A)** Violin plots showing the densities of individual immune cell phenotypes within the stromal and epithelial compartments of PDAC cores. The table below indicates the marker expression profile for each phenotype, with black squares with “+” denoting marker positivity and colours representing aggregate cell phenotypes employed in downstream analysis. **(B)** Forest plots of hazard ratios from univariate Cox regressions comparing high vs low densities of each phenotype. Solid red boxes indicate phenotypes significant in both univariate and multivariate models; dotted red boxes denote phenotypes significant in univariate analysis only.

### Increased T cell density is associated with greater overall survival

Previous studies indicated that T cells, B cells, and macrophages, and their specific subsets inform good or bad prognosis in PDAC,^36–39^ thus we sought to determine whether immune cell densities in PDAC tumour compartments correlated with OS. We used univariate and multivariate Cox regression analyses to compare 5-year OS in patients with immune cell densities categorized as high or low.

By univariate analysis, higher densities of epithelial CD16a+ cells were associated with an increased risk of death (HR: 2.4, 95% CI: 1.4–4.4, p<0.05), while higher densities of CD3+CD8– T cells in both the stroma and total tissue compartments were significantly associated with reduced risk of death (HR: 0.55, 95% CI: 0.31–0.97, p<0.05; HR: 0.42, 95% CI: 0.24–0.75, p<0.05, respectively) **(Figure 2B)**. These associations remained significant after adjusting for regional lymph node involvement by multivariate assessment (epithelial CD16a+: HR: 2.1, 95% CI: 1.1-3.9, p<0.05; stromal CD3+CD8-: HR: 0.47, 95% CI: 0.26-0.85, p<0.05); total CD3+CD8-: HR: 0.43, 95% CI: 0.24-0.80, p<0.05) **(Supplemental Table 3)**. Increased M1-like macrophage density was associated with decreased risk of death in univariate analysis (HR: 0.48, 95% CI: 0.26–0.89, p<0.05), but this association was lost in the multivariate model (HR: 0.55, 95% CI: 0.30-1.0. Thus, immune cell infiltration itself does not suggest improved prognosis: different subsets are variably prognostic and CD3+CD8– T cells may be a marker of improved OS in patients with PDAC.

### Combined CD3+CD8– and CD3+CD8+ T cells infiltration is associated with improved OS

Immune cells function as a coordinated network. We next hypothesized that the association of CD3+CD8-T cells with improved OS might reflect a role for these cells–which are likely helper T cells– in supporting anti-tumour immune functions, including that of cytotoxic (CD3+CD8+ T cells). Indeed, patients with high infiltration of both CD3+CD8- and CD3+CD8+ T cell subsets exhibit significantly improved OS compared to those low for both (HR: 0.33, 95% CI: 0.12–0.88, p<0.05). While high infiltration of either subset alone trended toward lengthened OS, neither reached statistical significance (CD3+CD8– only: HR: 0.45, 95% CI: 0.17-1.2, p =0.14; CD3+CD8+ only: HR 0.65, 95% CI: 0.14-3.0, p=0.88) **(Figure 3A)**. Notably, high infiltration of CD3+CD8+ T cells alone was uncommon (7% of patients) **(Figure 3B)**. These findings suggest that co-infiltration of helper and cytotoxic T cells might enable local generation of antitumour immunity.

**Figure 3.**
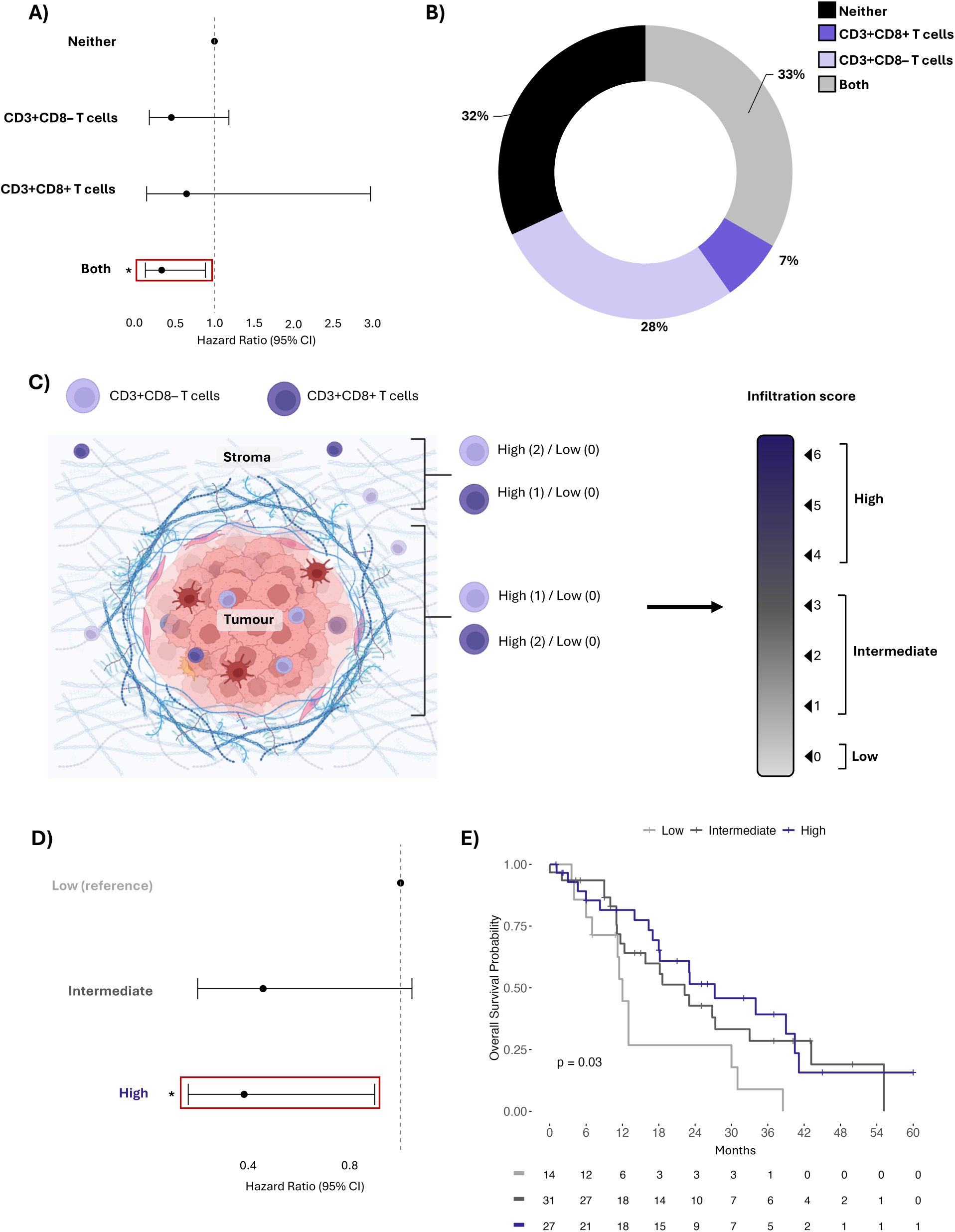
Combined CD3+CD8- and CD3+CD8+ T cell infiltration is associated with improved OS. **(A)** Forest plot showing hazard ratios and 95% confidence intervals from univariate Cox regression comparing patients categorized as high for CD3+CD8+ only, CD3+CD8-only, both, or neither, based on total cell density. **(B)** Pie chart representing the percentage of patients in each T cell group. **(C)** Schematic of the T cell infiltration score. Created in BioRender. Arseneau, R. (2025) https://BioRender.com/teuxamp **(D)** Forest plot showing hazard ratios and 95% confidence intervals from univariate Cox regression comparing patients based on infiltration score (low, intermediate, and high). Kaplan-Meier survival plot comparing the five-year OS among patients stratified by infiltration score.

#### High T cell infiltration score is predictive of longer OS

CD8+ cytotoxic T cell killing is contact-dependent, but helper T cells can function from a distance. To evaluate whether compartment-specific localization influences prognosis, we generated a T cell score **(Figure 3C)**. High T cell infiltration scores were associated with reduced risk of death and improved survival compared to low in both univariate and multivariate Cox regression models (HR: 0.39, 95% CI: 0.16-0.90, p<0.05) and Kaplan-Meier survival analysis (p<0.05) **(Figure 3D&E, Supplemental Table S3)**. Patients with intermediate T cell infiltration scores exhibited a trend towards decreased risk of death and lengthened OS compared to those with low scores, though the difference did not reach statistical significance (HR: 0.46, 95% CI: 0.20-1.0, p=0.066) **(Figure 3D&E)**. These findings suggest that the localization of T cell subsets with respect to tumour cells impact patient prognosis.

#### Higher peritumoral CD3+CD8– T cell density corresponds to decreased risk of death

PDAC tumours grow as cells around pancreatic ducts, dispersed between dense stroma throughout the pancreas; this is different from other solid tumours where groups of cancerous cells aggregate.^19,20^ Dichotomizing tumour versus stroma, as with other tumours, might therefore inadequately capture immune:tumour interactions in PDAC. To refine the assessment of spatial immune localization and its association with survival, we applied DBSCAN-based spatial clustering to segment tissue into intratumoural, peritumoural, and distal stromal regions and measure immune cell localization within them **(Figure 4A).** This approach enabled a more granular evaluation of immune cell positioning at the tumour-stroma interface.

**Figure 4.**
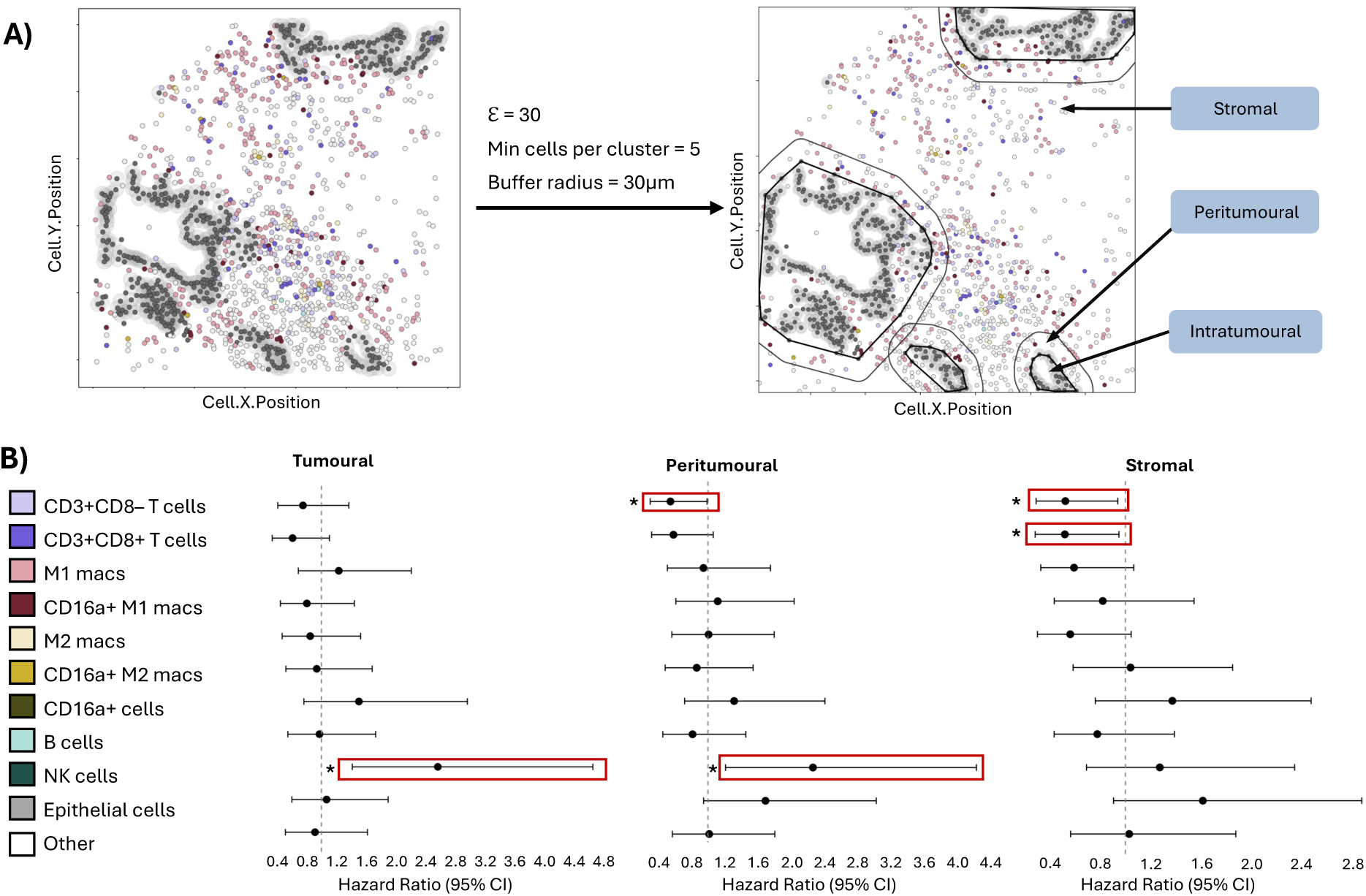
Immune cell proximity to the tumour predicts survival. **(A)** Schematic of DBSCAN-based clustering analysis. **(B)** Forest plots of hazard ratios from univariate Cox regressions comparing high vs low proportions of each phenotype in each region. Solid red boxes indicate phenotypes significant in both univariate and multivariate models; dotted red boxes denote phenotypes significant in univariate analysis only.

Within the intratumoural region, higher proportions of NK cells were associated with increased risk of death (HR: 2.6, 95% CI: 1.4-4.7, p<0.05) **(Figure 4B, Supplemental Table 3)**. Similarly, increased NK cell infiltration in the peritumoural space was also associated with worse survival (HR: 2.3, 95% CI: 1.2-4.2, p<0.05). In contrast, higher proportions of CD3+CD8-T cells in the peritumoural region were significantly associated with reduced risk of death (HR: 0.55, 95% CI: 0.30-0.99, p<0.05). Within the distal stroma, both CD3+CD8– and CD3+CD8+ T cells were associated with improved OS (HR: 0.53, 95% CI: 0.29-0.94, p<0.05; HR: 0.53, 95% CI: 0.29-0.95, p<0.05). Taken together, these findings suggest that T cell localization, particularly at the tumour-stroma interface, is predictive of prognosis in PDAC.

#### T cells and B cells colocalize in PDAC

Although T cell infiltration is often assessed for its prognostic capacity, effective immunity requires cooperation between immune cell subtypes. To determine whether additional cells might interact with T cells in the tumour environment, we performed Spearman’s correlation analysis on average immune cell densities across patients. Positive correlations were observed between CD3+CD8– and CD3+CD8+ T cells (R = 0.64, p<0.001), CD3+CD8– T cells and B cells (R = 0.71, p<0.001), and CD3+CD8+ T cells and B cells (R = 0.52, p<0.001) **(Figure 5A)**. These findings suggest that crosstalk may occur between lymphocytes, which may prompt formation of niches within the PDAC microenvironment.

**Figure 5.**
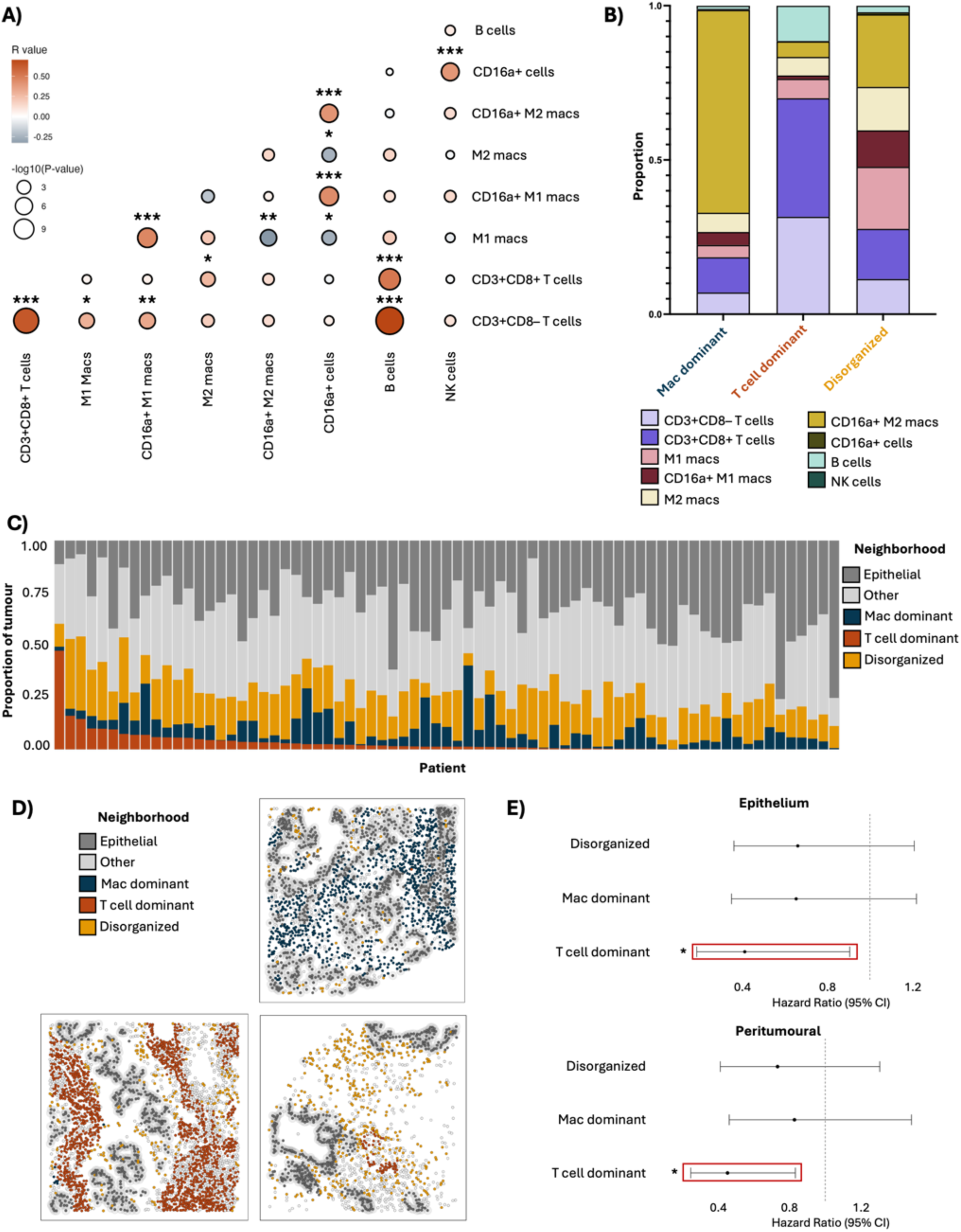
Co-infiltration and spatial organization of T cells and B cells is associated with improved patient outcomes in PDAC. **(A)** Co-infiltration of total cell densities in PDAC tumours, determined by Spearman’s correlation. Colour intensity indicates the strength of correlation, and dot size reflects the statistical significance (p-value). **(B)** Cell type proportions comprising the three immune neighbourhoods identified via spatial clustering. **(C)** Proportional representation of each immune neighbourhood in each of the 73 tumours assessed **(D)** Representative spatial neighbourhood maps demonstrating each neighbourhood subtype: Macrophage (Mac) dominant, T cell dominant, and Disorganized. **(E)** Forest plots of hazard ratios and 95% confidence intervals from univariate Cox regressions comparing “high” vs “low” proportions of each neighbourhood in the intratumoural region. Solid red box indicates neighbourhood significant in both univariate and multivariate models.

We next applied radius-based nearest neighbor clustering to ascertain potential interactions between immune cell types. With these, we identified three patterns of immune cell colocalization, which we termed “neighbourhoods”. The first was a *macrophage dominant* neighbourhood, composed primarily of CD16a+ M2-like macrophages, consistent with an immunosuppressive function **(Figure 5B).** The second was a *T cell dominant* neighbourhood, enriched for CD3+CD8– T cells, CD3+CD8+ T cells, and B cells, which may represent areas of organized adaptive immunity. The third was a *disorganized* neighbourhood, characterized by a heterogenous mix of infiltrating immune cells without a dominant cell type, suggestive of an unstructured (and perhaps inefficient) immune response.

The relative area represented by each neighbourhood varied between patients; however, in most tumours, the disorganized neighbourhood comprised the largest proportion of immune cells, followed by the macrophage dominant neighbourhood, and then the T cell dominant neighbourhood **(Figure 5C,D)**. The disorganized and macrophage dominant neighbourhoods were observed in all patients, while the T cell dominant neighbourhood was present in ∼90% of cases. Representative examples of each neighbourhood pattern are shown in **Figure 5D**.

To evaluate whether immune neighbourhoods were associated with patient outcomes, we performed Cox regression analyses comparing five-year OS in patients categorized as high or low for each neighbourhood. Having a higher density of the T cell dominant neighbourhood in the epithelium was associated with a decreased risk of death (HR: 0.41, 95% CI: 0.19-0.91, p<0.05). A higher proportion of the T cell dominant neighbourhood within the peritumoural region was associated with a decreased risk of death (HR: 0.45, 95% CI: 0.24-0.82, p<0.05) **(Figure 5E)**. These associations were retained after multivariate Cox regression analysis **(Supplementary Table 3)**. Neither the disorganized nor the macrophage dominant neighbourhoods were associated with OS. We conclude that the spatial co-localization of T cells and B cells proximal to the tumour is prognostically beneficial.

## Discussion

PDAC tumours have a characteristically dense and desmoplastic stroma. The relative lack of anti-tumour immune cells and highly immunosuppressive features are well-recognized.^4,5,40^ These observations have contributed to the prevailing view that PDAC may not be a suitable candidate for immunotherapy. However, recent studies reveal pockets of immune cell localization in PDAC,^7^ suggesting that, at least in some regions, PDAC tumours may remain immunologically susceptible. We profiled the immune landscape of 73 patients with PDAC, designing custom algorithms to assess the tumours’ unique ductal architecture. We found CD3+CD8-T cells to be associated with improved five-year OS, particularly when they localize to the tumour-stroma interface and when they coincide with CD3+CD8+ T cells in the tumour epithelium. T and B cell co-infiltration and co-localization were associated with the longest OS, supporting our conclusion that productive antitumour immunity results from the recruitment of cooperative immune cell types. This organization is consistent with that of the “T cell dominant” neighbourhood, where it would be expected that cytotoxic T cell immunity and antibody production could occur. This implies a susceptibility of PDAC to immune-mediated mechanisms and sets type I immunity as a goalpost for its future immunotherapy.

Immune profiling using IHC, single-cell mass cytometry, and flow cytometry has demonstrated that the extent and composition of T cell infiltration varies across patients with PDAC.^37,38,41–43^ Transcriptomic analyses identified distinct immunologic subtypes in PDAC, including those that are immune-rich, immunosuppressive, immune-exhausted, or immune-depleted, reflecting both the extent and functional state of immune infiltration.^6–10,38^ While transcriptomic subtypes reflect functional immune states, they do not fully capture the spatial heterogeneity observed within individual tumours. Recent studies demonstrate that tumours are not universally comprised of a single cell type or subregion.^7,44^ This regional variation, “sub-TMEs”, might reflect PDAC’s continued growth and diversification,^45,46^ and possibly a co-evolution with immunity and immune escape. We similarly observe a range of immune cell densities between patients, with CD3+CD8– T cells (likely CD4+ T cells), CD3+CD8+ T cells, and CD16a+ M2-like macrophages representing the dominant phenotypes in both the epithelial and stromal compartments. This is consistent with previous studies demonstrating that T cells are often the most abundant immune population in the PDAC stroma, even exceeding myeloid cells.^38,41,47^ With spatial assessments, we note that B cells are also included in these neighbourhoods, so the T cell populations are likely harbingers of more complex local immunity.

Higher densities of CD3+CD8– T cells in the stroma and T cell dominant neighbourhoods were associated with lengthened five-year OS. Due to limitations in panel size, we could not stain for CD4 directly, but infer these to be the CD3+CD4+ helper T cell subset. Likewise, an assessment of the features of CD4+ subsets is beyond the scope of our panel, but the co-localization of CD3+CD8-cells with B cells, M1 macrophages and CD3+CD8+ cells suggests their contribution to a type I immune response (likely associated with the Th1 helper cell subset). Since these cells associate with a survival benefit, we expect that they represent CD3+CD8–CD4+Foxp3– effector cells, which are known to correlate with improved survival in PDAC.^37^ CD4+ T helper cells promote anti-tumour immunity by orchestrating dendritic cell activation and licensing, facilitating CD8+ T cell recruitment, and supporting B cell maturation.^25^ Helper T cell localization near the tumour border may facilitate direct tumour cell killing through the release of cytotoxic cytokines (e.g. interferon-γ and tumour necrosis factor-α) or engagement of death receptors including TRAIL and FasL.^25,48,49^ Since these functions can occur even at a distance, our findings of benefit to OS of CD3+CD8– T cells in both the stroma and at the peritumoural regions still support a conclusion that these cells function as helpers. The prognostic significance of stromal CD3+CD8– T cells was further enhanced when CD3+CD8+ T cells were also present within the epithelium. Indeed, CD3+CD8+ T cells rarely infiltrate in the absence of CD3+CD8– T cells, underscoring that these helper populations may facilitate the recruitment or maintenance of the CD8+ populations in PDAC. CD3+CD8– T cells, CD3+CD8+ T cells, and B cells co-infiltrated and co-localized, comprising the primary cell types in the T cell-dominated neighbourhood. This spatial arrangement could reflect tertiary lymphoid structure (TLS)-like formations: organized groupings of immune cells that are associated with improved overall survival in PDAC^50–52^. TLSs support antigen presentation and adaptive immune activation; within them, interactions between B cells, CD4+ T cells, and CD8+ T cells – such as antigen presentation, co-stimulation, and cytokine-mediated signalling – promote coordinated immune responses.^18^ Although we did not have the necessary markers to definitively classify our T cell dominant neighbourhoods as TLSs, the composition of these two types of neighbourhoods is aligned, and their positioning at the tumour and peritumoural regions in patients with longer OS supports a productive anti-cancer immunity.

In addition to the T cell dominant neighbourhood, we defined two others: macrophage-dominant and disorganized. These may reflect immune subtypes previously described in PDAC. For example, the macrophage dominant neighbourhood, enriched for CD16a+ M2-like macrophages and exhibiting minimal T and B cell presence, may reflect an immunosuppressive environment consistent with previously defined “basal-like”, “squamous”, or perhaps even “immune escape”.^6,53,54^ The T cell dominant neighbourhood could reflect the previously defined “immune rich”, “reactive”, or “adaptive/T cell dominant” regions.^6–8^ Finally, the disorganized environment might reflect the “deserted” or “immune-exclusion/tumour dominant” tumour regions.^6,8^

PDAC has a distinctive histology in which tumour cells originate in the ducts and often spread as small, irregular clusters that are often interspersed within dense stroma.^19,20^ Existing approaches for spatial assessments were insufficient to capture this architecture, and methodological choices can influence interpretation. In our study, we first identified tumour clusters, then defined 30-µm borders around these clusters to capture both immune-immune and immune-tumour interactions occurring in their vicinity. This approach was designed to capture the histology of PDAC and improve biological relevance by reflecting the context in which immune activity at the tumour-stroma interface may be as relevant as direct tumour contact.^55,56^ Other studies using different spatial definitions (e.g. measuring immune cell proximity within 20-µm of each tumour cells) have reported associations between CD8+ T cells in direct contact with tumour cells and patient outcomes.^37^ This approach provides insight into direct tumour-immune interactions, but may not capture the broader immune context, including activity at the tumour-stroma interface. This may explain why CD4+ T cell proximity was not similarly associated with survival in those studies, despite their overall findings supporting the prognostic relevance of T cell infiltration.^37^ Our analysis, which used a different spatial framework, may reflect complementary aspects of PDAC histology and immune dynamics.

Most patients with PDAC are diagnosed at advanced stages, whereas our cohort consisted only of patients who underwent surgical resection (stage I-IIIa). Our findings thus reflect earlier-stage disease and may not fully capture the TME of metastatic PDAC. It is possible that patients with metastatic disease harbour a distinct TME.^45^ While we did not observe an association between tumour stage and immune contexture, it remains possible that tumours evolve toward becoming disorganized or immunologically-cold as they progress to advanced stages. Although functional data were beyond the scope of this work, our findings support future investigations and possible interventions to maximize the activity of the TLS-like/T cell dominant neighbourhoods in PDAC.

## Acknowledgements

We thank the patients who generously donated tissue and clinical information for this study, as well as the Nova Scotia Health Biobank for their contributions to sample and data collection. We are grateful to Craig’s Cause Pancreatic Cancer Society for their support. Dalhousie University and the Boudreau laboratory are situated in Mi’kmi’ki, the traditional and unceded territory of the Mi’kmaq. This work was supported by the Canadian Cancer Society’s Innovation (#705275) and JD Irving Atlantic Canada (#707195) grants to JEB. Clinical database creation was supported by a Nova Scotia Health Operating Grant and a Queen Elizabeth II Health Science Centre Foundation private donation research grant. RJA and JPM are trainees in the Cancer Research Training Program of the Beatrice Hunter Cancer Research Institute. Funds for RA and JPM are provided by GIVETOLIVE, by the Kilpatrick Trust through the Dalhousie Faculty of Medicine 2023 Graduate Studentship program, and by the Terry Fox MOHCCN Health Informatics and Data Scientist Awards. Funds for RA’s work is supported by the Rick Salsman Pancreatic Cancer Studentship Fund through the Dalhousie Faculty of Medicine 2025 Graduate Studentship program.

## Data availability

The data generated and code used in this study are available upon request from the corresponding author.

