## Supplementary information for "Peri-tumoural lymphocyte neighbourhoods predict longer survival in pancreatic ductal adenocarcinoma"

Supplemental information

**Supplemental Table 1. Clinical features of patients included in imaging and univariate/multivariate analysis.**

|  |  |  | **5-year OS** | | | **Associations with phenotype densities** | **Associations with neighborhood densities** |
| --- | --- | --- | --- | --- | --- | --- | --- |
| **Clinical variable** |  | Total n (%) | HR | 95% CI | p |  |  |
| **Age** |  |  |  |  |  |  |  |
|  | >65 | 43 (59%) | REF: Above 65 HR: 0.65 [0.36-1.2], p = 0.16 | | | None observed. | None observed. |
|  | <65 | 30 (41%) |  |  |  |  |  |
| **Sex** |  |  |  |  |  |  |  |
|  | Male | 40 (55%) | REF: Male HR: 0.57 [0.31-1.0], p=0.054 | | | Higher epithelial CD16a+ cell density in males (p<0.05). | None observed. |
|  | Female | 33 (45%) |  |  |  |  |  |
| **Stage*** |  |  |  |  |  |  |  |
|  | T1 | 2  (3%) | Stage I - Stage II: HR: 0.56 [0.7-4.3], p=0.95 Stage I - Stage III: HR: 0.67 [0.09-5.0], p=0.98 Stage II - Stage III: HR: 1.2 [0.66-2.2], p=0.94 | | | Higher density of “Other” cells in the epithelium in Stage II compared to stage III tumours (p<0.05).  Increased stromal CD16a+ M1-like macrophages and M1 macrophages in stage III compared to stage II (p<0.05). | None observed. |
|  | T2 | 24 (33%) |  |  |  |  |  |
|  | T3 | 45 (62%) |  |  |  |  |  |
|  | Not reported | 2 (3%) |  |  |  |  |  |
| **Grade** |  |  |  |  |  |  |  |
|  | 1 | 10 (14%) | Grade 1 vs Grade 2: HR: 0.66 [0.30-1.5], p=0.57 Grade 1 vs Grade 3: HR: 0.55 [0.22-1.4], p=0.41 Grade 2 vs Grade 3: HR: 0.83 [0.42-1.6], p=0.86 | | | Higher density of epithelial cells in the stromal compartment observed with increasing tumour grade (Grade 1 vs 3, p<0.05; Grade 2 vs 3, p<0.05). | None observed. |
|  | 2 | 46 (63%) |  |  |  |  |  |
|  | 3 | 17 (23%) |  |  |  |  |  |
|  | Not reported | 0 |  |  |  |  |  |
| **Regional lymph nodes** | | | | | | | |
|  | 1 | 12 (16%) | N0 vs N1: HR: 0.21 [0.05-0.89], p=0.085 N0 vs N2: HR: 0.16 [0.04-0.73], p=0.045 N1 vs N2: HR: 0.77 [0.40-1.5], p=0.73 | | | Decreasing epithelial cell density in the epithelium with increasing nodal involvement (N0 vs N1; N2 vs N3, p<0.05).  Increasing density of other cells with increasing node status (N1 vs N2, p<0.05). | None observed. |
|  | 2 | 44 (60%) |  |  |  |  |  |
|  | 3 | 15 (21%) |  |  |  |  |  |
|  | Not reported | 2 (3%) |  |  |  |  |  |

Abbreviations: overall survival (OS), hazard ratio (HR), confidence interval (CI), reference (REF).

*Numbers do not add to 100% due to rounding

**Supplemental Table 2 Cellular densities per tissue region.**

|  | Epithelium | | | Stroma | | | Total | | |
| --- | --- | --- | --- | --- | --- | --- | --- | --- | --- |
|  | Median | min | max | Median | min | max | Median | min | max |
| **CD3+CD8- T cells** |  |  |  |  |  |  |  |  |  |
| CD3+CD8– | 2.0 | 0 | 36.1 | 144.3 | 28.4 | 2905.0 | 115.2 | 19.6 | 2340.0 |
| CD3+CD8–CD16a+ | 0 | 0 | 1.1 | 0 | 0 | 6.5 | 0 | 0 | 5.5 |
| CD3+CD8–CD57+ | 0 | 0 | 0 | 0.4 | 0 | 4.0 | 0.3 | 0 | 3.0 |
| **Sum CD3+CD8– T cells** | 2.0 | 0 | 37.1 | 144.6 | 28.4 | 2909.0 | 115.2 | 20.1 | 2343.0 |
| **CD3+CD8+ T cells** |  |  |  |  |  |  |  |  |  |
| CD3+CD8+ | 10.6 | 0 | 214.0 | 233.0 | 20.8 | 1438.0 | 170.2 | 7.5 | 1167.0 |
| CD3+CD8+CD16a+ | 0 | 0 | 1.9 | 0.4 | 0 | 7.2 | 0.3 | 0 | 4.3 |
| CD3+CD8+CD57+ | 0 | 0 | 6.5 | 0.8 | 0 | 6.6 | 0.7 | 0 | 5.5 |
| **Sum CD3+CD8+ T cells** | 10.6 | 0 | 216.1 | 234.4 | 24.9 | 1446.0 | 176.4 | 8.0 | 1173 |
| **NK cells** |  |  |  |  |  |  |  |  |  |
| CD57+CD16a- | 0 | 0 | 1.1 | 0 | 0 | 2.3 | 0 | 0 | 1.9 |
| CD57+CD16a+ | 0 | 0 | 7.5 | 0.2 | 0 | 2.7 | 0.2 | 0 | 1.7 |
| **Sum NK cells** | 0 | 0 | 7.5 | 0.4 | 0 | 4.1 | 0.3 | 0 | 1.9 |
| M2 macs | 2.0 | 0 | 38.8 | 228.9 | 31.5 | 623.5 | 179 | 13.0 | 499.3 |
| CD16a+ M1 macs | 3.8 | 0 | 37.7 | 120.5 | 11.7 | 899.3 | 97.84 | 8.6 | 162.9 |
| M2 macs | 1.9 | 0 | 34.0 | 156.9 | 12.7 | 519.0 | 117.8 | 9.1 | 417.7 |
| CD16a+ M2 macs | 11.3 | 0 | 280.1 | 569.1 | 45.2 | 4602.0 | 396.0 | 38.1 | 2006.0 |
| CD16a+ cells | 0 | 0 | 7.7 | 7.3 | 1.0 | 52.6 | 5.7 | 0.7117 | 34.5 |
| B cells | 0 | 0 | 11.1 | 8.8 | 0 | 1138.0 | 5.3 | 0 | 915.9 |
| Epithelial cells | 5214.0 | 2377.0 | 8678.0 | 266.1 | 43.6 | 1319.0 | 1467.0 | 231.5 | 4167.0 |
| Other | 4.4 | 0 | 134.0 | 2343.0 | 1458.0 | 4278.0 | 1812.0 | 406.7 | 3117.0 |

Cell types indicated were assessed independently in initial analysis. Thereafter, CD3+CD8-, CD3+CD8+ or NK cell subpopulations were summed, as shown in grey lines. Numbers indicate cells/mm^2^.

**Supplemental Table 3. Multivariate Cox Regressions**

| **Variable** | Region | HR | 95% CI | p-value |
| --- | --- | --- | --- | --- |
| CD16a+ cells | Epithelium | 2.1 | 1.1-3.9 | 0.017 |
| CD3+CD8- T cells | Stroma | 0.47 | 0.26-0.86 | 0.014 |
|  | Total | 0.43 | 0.24-0.80 | 0.0070 |
|  | Peritumoural | 0.54 | 0.29-0.99 | 0.046 |
|  | Remaining stroma | 0.55 | 0.30-0.99 | 0.048 |
| CD3+CD8+ T cells | Remaining stroma | 0.47 | 0.25-0.88 | 0.019 |
| M1-like macrophages | Total | 0.55 | 0.30-1.0 | 0.069 |
| NK cells | Tumoural | 2.4 | 1.2-4.7 | 0.012 |
|  | Peritumoural | 2.2 | 1.2-4.1 | 0.012 |
| T cell dominant | Epithelium | 0.22 | 0.14-0.78 | 0.011 |
|  | Peritumoural | 0.40 | 0.10-0.78 | 0.0070 |

Abbreviations: hazard ratio (HR), confidence interval (CI). Analyses have been adjusted for lymph node involvement. Only those comparisons with significant p-values in univariate analyses are displayed.
